# National Norms and Psychometrics for the Pubertal Development Scale

**DOI:** 10.64898/2026.03.25.26349307

**Authors:** Yinuo Liu, Andrea E. Bonny, Guillermo Perez Algorta, Eric A. Youngstrom

**Affiliations:** Nationwide Children’s Hospital; The Ohio State University; Lancaster University

**Keywords:** Puberty, Assessment, Continuous Norm, Cross-Informant Agreement, Psychometrics

## Abstract

**Introduction:** The Pubertal Development Scale (PDS) is widely used for puberty assessment, yet prior investigations have been confined to research samples. This study examined PDS psychometrics and established continuous norms in a nationally representative U.S. sample.

**Methods:** We analyzed parent-report data for children aged 6-18 (*N*=2251), youth self-report data aged 12-18 (*N*=1006), and a retest sample of parents (*N*=171) and youth (*N*=90). Factor structure, internal consistency, measurement precision, test-retest reliability, and invariance were assessed. Cross-informant agreement was evaluated with intraclass correlation coefficients and Bland-Altman plots. Age– and sex-adjusted norms were established using Generalized Additive Models for Location, Scale, and Shape. Associations between pubertal timing and clinical symptoms were examined via multiple linear regression.

**Results:** The PDS demonstrated one latent factor after accounting for limited score variability among prepubertal children. Internal consistency was acceptable-to-good (Cronbach’s *α*=.77-.89; McDonald’s *ω*=.78-.90), conditional reliability exceeded .80 across approximately 13 of 15 possible scores, and test-retest reliability was excellent (Pearson’s *r*=.90-.96; ICC(2,1)=.89-.96). Differential item functioning across racial and parental education subgroups had negligible impact on total scores. Cross-informant agreement was excellent: ICC(2,1)=.87-.88. PDS scores increased nonlinearly with age, with girls showing earlier onset, faster progression, and greater convergence toward completion by late adolescence. Pubertal timing predicted parent-reported child symptoms of depression, mania, and sleep disturbance, beyond chronological age and sex.

**Discussion:** The PDS shows strong psychometrics in a nationally representative sample, supporting its utility in research and clinical contexts. The national norms provide empirical benchmarks for score interpretation, strengthening the PDS’s value as a pre-screener for atypical pubertal timing.

## Introduction

Puberty, the process of physical development to sexual maturity, is characterized by significant biological, hormonal, and mental changes (Breehl & Caban, 2018). Early or delayed puberty could have important implications on adolescent well-being. Early pubertal timing is associated with increased mental and behavioral health problems in both sexes, with a meta-analysis indicating a small magnitude of effect (Ullsperger & Nikolas, 2017): boys who mature early showed elevated risks of substance use (Patton et al., 2004) and delinquency (Williams & Dunlop, 1999), while girls who mature early experience more depression, anxiety, eating problems, and substance use (Angold et al., 1998; Kaltiala-Heino et al., 2003; Knight et al., 2021). Delayed puberty may signal underlying chronic illness or nutritional deficiencies, cause emotional distress, and impair physical growth (Palmert & Dunkel, 2012). Accurate assessment of puberty is therefore crucial for researchers and clinicians to identify atypical pubertal timing and examine adolescent outcomes.

The Pubertal Development Scale (PDS) is a self– and parent-report measure widely used in research and clinic settings to assess puberty (Petersen et al., 1988). It asks the respondent to estimate the level of completion of key pubertal events, including height spurt, body hair growth, skin changes, and sex-specific indicators. Originally developed by Petersen et al. as a self-report instrument for adolescents in sixth grade and above, PDS was later adapted to include a parent-report version, with both versions demonstrating good internal consistency and test-retest reliability in research samples (Koopman-Verhoeff et al., 2020). Compared to the clinician-administered Tanner staging, the PDS does not require trained specialists to conduct a physical examination while still showing moderate to good agreement with clinician judgements (Pan et al., 2025). Accordingly, it can be administered by any provider, including psychologists and other non-physician clinicians who lack the scope of practice for a physical exam, making it a practical alternative in behavioral health settings where puberty-related questions arise but genital or breast examination is not feasible or appropriate. This ease of administration and the lack of a fee for use have contributed to its popularity. The PDS has been translated into Chinese, Korean, and Portuguese, with reliability and validity maintained across these languages and cultural contexts (Chan et al., 2010; Kim et al., 2025; Pompéia et al., 2019). As a low-cost, non-invasive instrument, the PDS has also been adopted in large-scale studies such as the Adolescent Brain Cognitive Development (ABCD) Study (Beltz et al., 2026; Cheng et al., 2021).

Despite this demonstrated validity and widespread adoption, the psychometric foundations of the PDS have not been thoroughly examined. Although the scale has been adapted to include a parent-report version and extended to serve a broader age range than the original middle-school sample, its unidimensional factor structure has not been formally evaluated in the expanded use case. Without this evidence, the validity of summing or averaging PDS items rests on an untested assumption. Reliability evidence is similarly incomplete: internal consistency has typically been reported using Cronbach’s alpha alone, which yields a single global estimate without revealing where along the score range the scale is more or less precise, and test-retest estimates have been derived from small, non-representative samples (Koopman-Verhoeff et al., 2020). Through more advanced measurement methods such as Confirmatory Factor Analysis (CFA) and Item Response Theory (IRT), together with a representative national cohort, we can better evaluate the scale’s latent structure, conditional precision, temporal stability, and measurement equivalence across the age groups, sexes, and demographic subpopulations in which the PDS is routinely deployed.

Compounding these psychometric uncertainties, nationally representative normative benchmarks for the PDS do not yet exist. The scale’s initial norms were established by Petersen et al. (Petersen et al., 1988) with 355 adolescents aged 11-14 years, and subsequent U.S. studies have been constrained by small sample sizes (Brooks-Gunn et al., 1987; Koopman-Verhoeff et al., 2020) or incomplete demographic representation (Cheng et al., 2021). Without age– and sex-referenced benchmarks derived from a representative population, a raw PDS score carries limited information about a child’s pubertal development relative to same age and sex peers, leaving researchers without a shared standard for cross-study comparisons and clinicians without an empirical basis for identifying atypical pubertal timing. Establishing national norms would directly address this gap, transforming PDS scores from descriptive totals or averages into clinically interpretable metrics of pubertal timing.

Norming the PDS requires careful consideration of sex and age effects, as puberty is characterized by marked sex differences and peak development at certain ages. Capturing the nonlinear trajectories of pubertal development calls for a continuous norming approach (Lenhard et al., 2018), such as the Generalized Additive Models for Location, Scale, and Shape (GAMLSS) (Rigby & Stasinopoulos, 2005). The GAMLSS approach enables modeling of all features of the score distribution, including its central tendency, spread, and shape, while supporting hundreds of candidate score distributions. Unlike traditional norming methods that assume scores are normally distributed and assign children to discrete age categories, GAMLSS generates smooth norms that vary continuously with age, which offers considerably greater flexibility. In simulations, GAMLSS outperformed traditional and polynomial methods and achieved higher norming accuracy with smaller samples (Frazier et al., 2026). These strengths have made GAMLSS increasingly applied in the norming of psychological tests (R. A. Kenny et al., 2013; Liu et al., 2025; Rubio-Guerra et al., 2025). Its capacity to model score parameters as continuous functions of age is particularly well-suited for capturing pubertal development, where the population average and between-person heterogeneity, reflected by the center and spread of PDS scores, are both of substantive interest. The resulting continuous norms can transform raw PDS scores into age– and sex-referenced indices, broadening their utility across research, clinical, and epidemiological contexts.

One important application of continuous PDS norms is to investigate puberty-outcome associations, in which the strength of existing evidence varies considerably by domain. Depression is the most extensively studied outcome: a large Danish population-based cohort found that earlier timing and faster tempo of pubertal milestones were associated with depressive symptoms and clinically diagnosed depression in both sexes (Benson et al., 2026), though a psychosocially-informed analysis of another national cohort found that pubertal timing itself explained no significant variance once childhood emotional problems and peripubertal parental and body-image context were accounted for (Duchesne et al., 2026), suggesting that timing’s association with depression may operate largely through, rather than independent of, psychosocial pathways. Puberty’s relationship to sleep disturbance is also well-characterized, with pubertal onset driving a slower accumulation of homeostatic sleep pressure and a melatonin-mediated circadian phase delay (Kansagra, 2020). Its relationship to ADHD hyperactivity and inattention symptoms is less settled, with direction and magnitude varying by whether timing, tempo, or status is examined and by informant (Porter et al., 2024). The association with manic symptoms is the least established, with existing literature remaining largely theoretical and the interaction between gonadal hormone changes and manic symptoms receiving comparatively little empirical attention relative to other outcomes (Colic et al., 2025). Much of this literature has been constrained by small (Alloy et al., 2016) or demographically restricted samples (Knight et al., 2021) and broad categorical classifications of early, on-time, and late puberty (Angold et al., 1998; Sun et al., 2017), which limits the generalizability of those findings.

A large, nationally representative U.S. sample with clinical symptom measures collected from both parents and children offers a stronger basis for examining the associations between atypical pubertal timing and mental and behavioral health outcomes. In particular, a continuous, norm-referenced pubertal timing index allows for a rigorous test of whether atypical development carries predictive value for clinical symptoms beyond age and sex, and whether this relationship holds across informant perspectives. It also provides an opportunity to examine pubertal associations with child mental and behavioral problems independent of referral and treatment-seeking patterns and their potential confounding impact (Berkson, 1946).

The present study addresses these psychometric and normative gaps through a systematic evaluation of the PDS in a nationally representative U.S. sample. We examine the scale’s psychometric properties using modern measurement methods, establish continuous national norms using the GAMLSS approach, and leverage these norms to investigate associations between pubertal timing and child clinical symptoms. By including both parent– and youth self-report data from a large, demographically diverse national sample, we aim to strengthen the empirical foundation of the PDS and advance its utility as an evidence-based tool for pubertal assessment in research, pediatric practice, and clinical psychology settings where a norm-referenced, non-invasive index of pubertal timing can inform case conceptualization and referral decisions.

## Methods

### Participants

We analyzed a deidentified, nationally representative survey data of U.S. families, in which both parents and their children completed measures independently. The full sample comprised 2,251 parent-reported assessments of children aged 6-18, of whom 1,006 children aged 12-18 also completed youth self-report measures, thus forming parent-child dyads. The sample was representative of the U.S. population on key demographics (child sex, race, ethnicity) and aligned closely with 2023 American Community Survey (ACS) estimates. Sample demographics were summarized along with 2023 ACS estimates in Table 1.

**Table 1.** Participant demographic characteristics.

|  | Parent Sample<br>(N = 2251) | Youth Sample<br>(N = 1006) | 2023 ACS<sup>a</sup> |
| --- | --- | --- | --- |
| Child age |  |  |  |
| M (SD) | 11.74 (3.40) | 14.43 (1.65) |  |
| 6-7 years | 15.5% |  |  |
| 8-9 years | 14.7% |  |  |
| 10-11 years | 14.3% |  |  |
| 12-13 years | 19.3% | 34.0% |  |
| 14-15 years | 19.9% | 35.2% |  |
| 16-18 years | 16.4% | 30.8% |  |
| Child sex |  |  |  |
| Female | 47.5% | 49.7% | 50.5% |
| Male | 52.5% | 50.3% | 49.5% |
| Race |  |  |  |
| White | 75.3% | 74.8% | 60.5% |
| Black or African American | 8.8% | 9.7% | 12.1% |
| Asian | 3.7% | 1.9% | 6.0% |
| American Indian or Alaska Native | 1.7% | 0.8% | 1.0% |
| Native Hawaiian or Pacific Islander | 0.3% | 0.2% | 0.2% |
| Some other race | 4.7% | 3.3% | 7.4% |
| Multiple races selected | 4.0% | 8.9% | 12.8% |
| Ethnicity |  |  |  |
| Hispanic, Latino, or Spanish Origin | 16.4% | 14.3% | 19.4% |
| Not Hispanic, Latino, or Spanish Origin | 83.6% | 85.7% | 80.6% |
| Parent/guardian education |  |  |  |
| High school graduate or higher | 97.6% | 97.7% | 89.8% |
| Bachelor's degree or higher | 47.5% | 48.7% | 36.2% |
| Median Household Income | \$70,000 - \$79,999 | \$70,000 - \$79,999 | \$77,719 |

<sup>a</sup>Data from 2023 American Community Survey (ACS) 1-year estimates.

In the parent-report data, children were on average 11.74 years old (range: 6-18, *SD*=3.40), with 47.5% of children being female. The majority of adolescents (ages 12-18) subsequently formed parent-child dyads with a youth self-report component (*n*=1,006). Based on parent report, 75.3% of children were White, 8.8% Black or African American, 3.7% Asian, and 16.4% Hispanic, Latino, or Spanish Origin. Nearly all parents (97.6%) had obtained a high school diploma, and 47.5% had obtained a bachelor’s degree or higher. The median household income was between $70,000 and $79,999 annually.

In the youth self-report data, children were on average 14.43 years old (range: 12-18, *SD*=1.65) with 49.7% female. Based on youth self-identification, 74.8% were White, 9.7% Black, 1.9% Asian, and 14.3% Hispanic. Because the youths and parents were linked as dyads, the household characteristics are similar as for the full parent sample: most parents (97.7%) had a high school diploma, and 48.7% of parents had a bachelor’s degree or higher. The median household income was also between $70,000 and $79,999 annually.

A subset of parents (*N*=171) and youth (*N*=90) was recontacted to complete the scale battery a second time. The average retest interval was 18.90 days (*SD*=6.45) for parents and 19.39 days (*SD*=7.78) for youth. The retest sample characteristics were highly similar to those of the full sample (see Supplemental Table S1 for details), indicating that participant composition did not differ meaningfully between the two waves, which supports the examination of test-retest reliability.

### Measures

The *Pubertal Development Scale* (PDS) is a five-item measure of pubertal status (Petersen et al., 1988). The first three items, covering height spurt, body hair growth, and skin changes, are identical for all children. The remaining two items are sex-specific: boys are asked about voice deepening and facial hair growth, and girls about breast development and menarche. Every item except menarche is rated on a 4-point Likert-type scale (1 = not yet started, 2 = barely started, 3 = definitely started, 4 = seems complete), whereas menarche is reported dichotomously and scored on the same metric (1 = no, 4 = yes). Prior studies have commonly averaged the five items, yielding scores from 1 to 4. To avoid this extra calculation and to preserve the granularity of score changes, we instead summed the items into a PDS total score ranging from 5 to 20 for all modeling and norms reported here. Percentile curves and reference tables rescaled to the 1-4 average-score metric are provided in Supplemental Figure S2 for parent-report data and S3 for youth self-report data.

#### Behavioral Correlates of Change in Pubertal Status

Parents and youth also completed the following measures of child mental and behavioral health. The *Very Quick Inventory of Depressive Symptomatology* (VQIDS) is a 5-item measure of core depression symptoms, including sad mood, self-outlook, involvement, fatigue, and concentration/decision-making, derived from the well-validated 16-item Quick Inventory of Depressive Symptomatology (Rush et al., 2003). The VQIDS demonstrates a single-factor structure, acceptable internal consistency, and sensitivity to symptom change comparable to that of the full-length version (De La Garza et al., 2017). The *Child Mania Rating Scale* (CMRS) assesses the frequency and severity of manic symptoms in children and adolescents, demonstrating high internal consistency and robust convergence with clinician-administered gold-standard mania scales (Pavuluri et al., 2006). Parents completed the full-length 21-item CMRS, and youth completed the shortened 10-item version. The *Vanderbilt Attention Deficit/Hyperactivity Disorder Parent Rating Scale* (VADPRS) screens for ADHD symptoms and comorbid conditions and rates their severity (Wolraich et al., 2003). We administered its hyperactivity and inattention symptom subscales to both parents and youth. Finally, the *Patient-Reported Outcomes Measurement Information System* (PROMIS) is a set of standardized measures developed by the National Institutes of Health to assess a range of physical, mental, and social health symptoms (Cella et al., 2010). We used the PROMIS Sleep-Related Impairment scale (Forrest et al., 2018) to evaluate children’s sleep disturbance. For scales without both parent-report and youth self-report versions, we adapted the available version so that each construct could be assessed by both informants.

### Procedure

This study was approved by the Institutional Review Board at Nationwide Children’s Hospital. We contracted with the survey agency YouGov to administer the PDS along with other clinical scales to parents and children. YouGov is a research and data analytics company that has a global panel of over 29 million registered members with a significant portion in the U.S., which enables the company to conduct national surveys. YouGov collected and cleaned all responses in late 2025, paired parent and child data to create dyads, and applied stratification based on key demographics (child sex, race, ethnicity, and parent education level) to align with the 2023 ACS 1-year estimates, resulting in nationally representative samples.

### Statistical Analyses

Analyses were stratified by child sex and informant type (parent-report vs. youth self-report) for three reasons: (1) pubertal processes differ by sex, (2) the PDS contains sex-specific items, and (3) parent and child perception of pubertal development could diverge.

#### Factor Structure

The PDS is designed to measure one underlying dimension of pubertal development, so we tested a one-factor measurement model with confirmatory factor analysis (CFA) using the lavaan package (Rosseel, 2012). Models were estimated from polychoric correlations with the robust weighted least squares mean– and variance-adjusted (WLSMV) estimator under the theta parameterization. The analytic sample contained no missing values on PDS items. Because the PDS was originally designed as a self-report instrument for youth aged 12 and above, floor effects from low puberty scores were anticipated in the parent-report sample of younger and prepubertal children (age range: 6-11) and had the potential to distort CFA model fit. We therefore fitted the one-factor model in the full parent-report sample and separately within the 6-11 and 12-18 age bands. The youth self-report sample covers ages 12-18 only, so no age-band split applied. Model fit was evaluated with the scaled *χ*^2^ test and scaled fit indices: the comparative fit index (CFI), the root mean square error of approximation (RMSEA) with its 90% confidence interval, and the standardized root mean square residual (SRMR).

#### Internal Consistency and Measurement Precision

To establish the psychometric reliability of the PDS, we evaluated its internal consistency using Cronbach’s *α* (Cronbach, 1951) and McDonald’s *ω* (McDonald, 1999), with the assumption of unidimensionality.

To examine measurement precision across the observed score range, we fitted Item Response Theory (IRT) models to each PDS version and applied Expected A Posteriori (EAP) scoring to map the latent trait (θ) onto the observed score metric (Bock & Mislevy, 1982). Items with 4-level responses were modeled with Graded Response Model, and the dichotomous menarche item was modeled with a two-parameter logistic model. Conditional reliability was then estimated and plotted across the full score range.

#### Test-Retest Reliability

Test-retest reliability of PDS total scores between the original and retest administrations was examined using Pearson’s *r* and Intraclass Correlation Coefficient (ICC). Pearson’s *r* indexes the relative consistency of participants’ rank ordering across the two occasions. ICC, specified as a two-way random effects model with absolute agreement and single unit of analysis (ICC(2,1)), penalizes any systematic mean-level change between test and retest and provides a stricter index of absolute score stability over time. ICC values range from 0 to 1, where 0 indicates no agreement and 1 indicates perfect agreement, with thresholds defined as <0.2 poor, 0.2-0.4 fair, 0.4-0.6 moderate, 0.6-0.8 good, 0.8-1.0 excellent (Landis & Koch, 1977).

#### Measurement Invariance

To determine whether the PDS items function equivalently across demographic groups, we tested differential item functioning (DIF) within an IRT framework. Two grouping variables were examined: child race (White vs. Non-White) and parental education (bachelor’s degree or higher vs. less than a bachelor’s degree), because our sample characteristics deviate most from the 2023 ACS estimates on these two demographic variables. The parent-report sample was split by age band (6-11 and 12-18) to distinguish DIF from developmental variation in pubertal status.

DIF was tested with likelihood ratio tests comparing a fully invariant multiple-group baseline model against models with freely estimated slope and intercept parameters of one item (Tay et al., 2015). Anchor items were selected by iterative purification, and *p* values were adjusted with false discovery rate correction (Benjamini & Hochberg, 1995). Items showing significant DIF were further examined for the direction and consistency of the DIF effect (uniform vs. nonuniform). Because item-level DIF does not necessarily bias total scores, we also evaluated Differential Test Functioning (DTF) to determine whether any item-level DIF produced meaningful differences in expected PDS total scores. We computed signed DTF (sDTF; systematic unidirectional bias) and unsigned DTF (uDTF; total discrepancy regardless of direction) using the focal group’s ability distribution as the weighting function.

#### Cross-Informant Agreement

We calculated ICC(2,1) (as described above) for parent-child dyads to assess cross-informant agreement. To visualize the agreement between parent– and self-report PDS scores, we created Bland-Altman plots (Bland & Altman, 1986) depicting the difference between informants (y-axis) against their mean scores (x-axis). Mean difference and limits of agreement were examined.

#### Regression-Based Continuous Norming

We established age– and sex-specific national norms for parent-report and self-report PDS with GAMLSS. Guided by Timmerman et al. (Timmerman et al., 2021, 2025), we compared multiple candidate distributions and selected the best-fitting model based on global and local fitness indices, including Bayesian Information Criterion (BIC) values (see Supplemental Table S2), worm plots [Buuren and Fredriks (2001); see Supplemental Figure S1], and Q-statistics. The Box-Cox Cole-Green (BCCG) distribution, with three parameters (*μ*, *σ*, *ν*), best fitted parent-report PDS data. Though the discrete Beta-Binomial (BB) distribution best fitted the youth self-report data, we selected the second-best BCCG distribution to generate smooth percentile curves and to enhance the modeling consistency across informant types. All GAMLSS models used P-spline smoothers to model age-dependent changes in the median (*μ*) and variation (*σ*). The skewness parameter (*ν*) was modeled as a P-spline function of age for parent-report boys and a linear function of age for parent-report girls, and held constant for youth self-report scores of both sexes. For each model, smooth percentile curves (5^th^, 10^th^, 25^th^, 50^th^, 75^th^, 90^th^, 95^th^) were estimated and plotted, and percentile tables were created for clinical reference. As recommended by Blagus et al. (Blagus et al., 2025), we developed a Shiny app to facilitate the conversion between PDS total scores and percentiles based on the informant type, child’s age, and sex (https://yinuoliu.shinyapps.io/ShinyApp/). The app is included here for demonstration purposes and is currently being updated with additional features.

#### Correlates of Norm-Referenced Pubertal Timing

To isolate the independent contribution of pubertal development to clinical symptom severity, we calculated each participant’s age– and sex-adjusted PDS percentile, which indexes pubertal timing relative to same-age, same-sex peers. Because clinical scales differ in their units and score ranges, we applied *z*-score standardization (*M*=0, *SD*=1) to all clinical scale total scores and to the PDS percentile prior to analyses. We then estimated multiple linear regression models predicting each of five clinical outcomes (depression, mania, hyperactivity, inattention, and sleep disturbance) from child sex, age, pubertal timing based on normative percentile, age-by-pubertal-timing interaction, and sex-by-pubertal-timing interaction, with *p*-values adjusted for false discovery rate. Unlike other analyses stratified by child sex and informant type, the regression models pooled data across sex within each informant sample.

## Results

### Factor Structure

In the full parent-report sample (ages 6-18), the one-factor PDS model showed excellent fit for boys (*χ*^2^(5)=13.86, *p*=.017, CFI=1.000, RMSEA=.039, SRMR=.013) and adequate-to-good fit for girls (*χ*^2^(5)=71.80, *p*<.001, CFI=.997, RMSEA=.112, SRMR=.025) (see Table 2). The RMSEA of the girls’ one-factor model exceeded conventional cutoffs. RMSEA is susceptible to inflation when a scale comprises few items, as the limited degrees of freedom can produce artificially elevated values (D. A. Kenny & McCoach, 2003), whereas SRMR does not share this sensitivity and may therefore provide a more reliable indicator of model fit. This elevated RMSEA of girl’s CFA model may also be attributable to floor effects among pre-pubertal children, which restricted item-response variability and distorted model fit indices. We tested this floor-effect interpretation by fitting one-factor CFA models separately within each age band of the parent-report sample. In the 6-11 age band, fit indices were markedly poorer for both boys (CFI=.981, RMSEA=.111, SRMR=.065) and girls (CFI=.975, RMSEA=.132, SRMR=.086), consistent with restricted variability in pubertal status among younger children. In contrast, within the 12-18 age band, model fit improved substantially for both boys (*χ*^2^(5)=11.05, *p*=.050, CFI=.999, RMSEA=.043, SRMR=.017) and girls (*χ*^2^(5)=6.32, *p*=.276, CFI=.999, RMSEA=.021, SRMR=.028).

**Table 2.** Confirmatory factor analysis fit indices for the one-factor PDS model by informant, child sex, and age band.

| Sex | Age | *n* | $\chi^2$ (*df*) | *p* | CFI | RMSEA [90% CI] | SRMR |
| --- | --- | --- | --- | --- | --- | --- | --- |
| Parent Report |  |  |  |  |  |  |  |
| Boys | 6-18 | 1,182 | 13.86 (5) | .017 | 1.000 | .039 [.015, .064] | .013 |
|  | 6-11 | 539 | 38.16 (5) | <.001 | .981 | .111 [.080, .145] | .065 |
|  | 12-18 | 643 | 11.05 (5) | .050 | .999 | .043 [.000, .078] | .017 |
| Girls | 6-18 | 1,069 | 71.80 (5) | <.001 | .997 | .112 [.090, .135] | .025 |
|  | 6-11 | 461 | 45.36 (5) | <.001 | .975 | .132 [.099, .169] | .086 |
|  | 12-18 | 608 | 6.32 (5) | .276 | .999 | .021 [.000, .063] | .028 |
| Youth Self-Report |  |  |  |  |  |  |  |
| Boys | 12-18 | 506 | 9.00 (5) | .109 | .998 | .040 [.000, .081] | .018 |
| Girls | 12-18 | 500 | 7.82 (5) | .166 | .999 | .034 [.000, .077] | .035 |

*Note.* All models specify a single latent pubertal development factor indicated by the five PDS items, estimated separately by informant, child sex, and age band because the PDS contains sex-specific items. CFI = comparative fit index; RMSEA = root mean square error of approximation; SRMR = standardized root mean square residual.

The one-factor model also fit well in the youth self-report sample (ages 12-18) regardless of child sex (boys: *χ*^2^(5)=9.00, *p*=.109, CFI=.998, RMSEA=.040, SRMR=.018; girls: *χ*^2^(5)=7.82, *p*=.166, CFI=.999, RMSEA=.034, SRMR=.035). Together, these results confirm that the PDS is best represented as a unidimensional pubertal development factor for both informants and sexes, once floor effects from limited pubertal development among children under 12 are accounted for by restricting the parent-report CFA to an age-appropriate group.

### Internal Consistency and Measurement Precision

For parent-report PDS, boys and girls demonstrated good internal consistency (Cronbach’s *α*=0.89 for both sexes; McDonald’s *ω*=0.89 for boys and 0.90 for girls; see Table 3). For self-report PDS, internal consistency was good for boys (*α*=0.84, *ω*=0.84) and acceptable for girls (*α*=0.77; *ω*=0.78). Both formats of PDS consistently measure the latent construct of puberty, but parent-report PDS showed higher consistency.

**Table 3.**
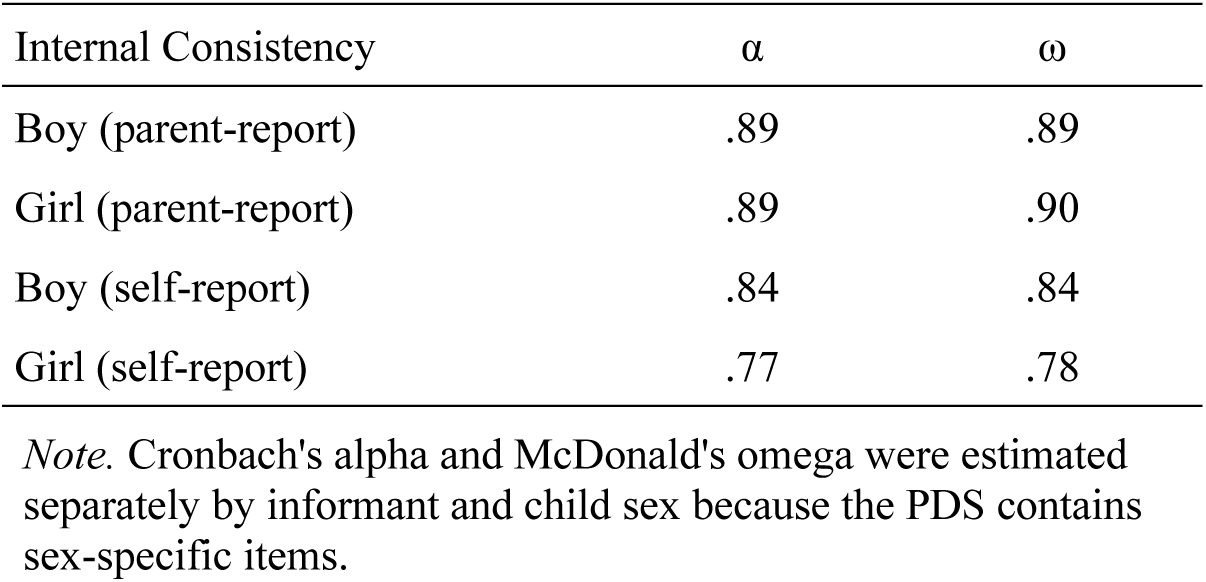
PDS internal consistency by informant and child sex.

Since *α* and *ω* represent average precision over the entire sample, we complemented them with conditional reliability across the observed score range (Figure 1). For parent-report PDS, reliability was greater than .80 at the score range of 7.2-20 for boys (Figure 1A) and 6.4-19.5 for girls (Figure 1B). Precision was lowest at the floor, consistent with the restricted item-response variability observed in the CFA models for younger, prepubertal children

**Figure 1.**
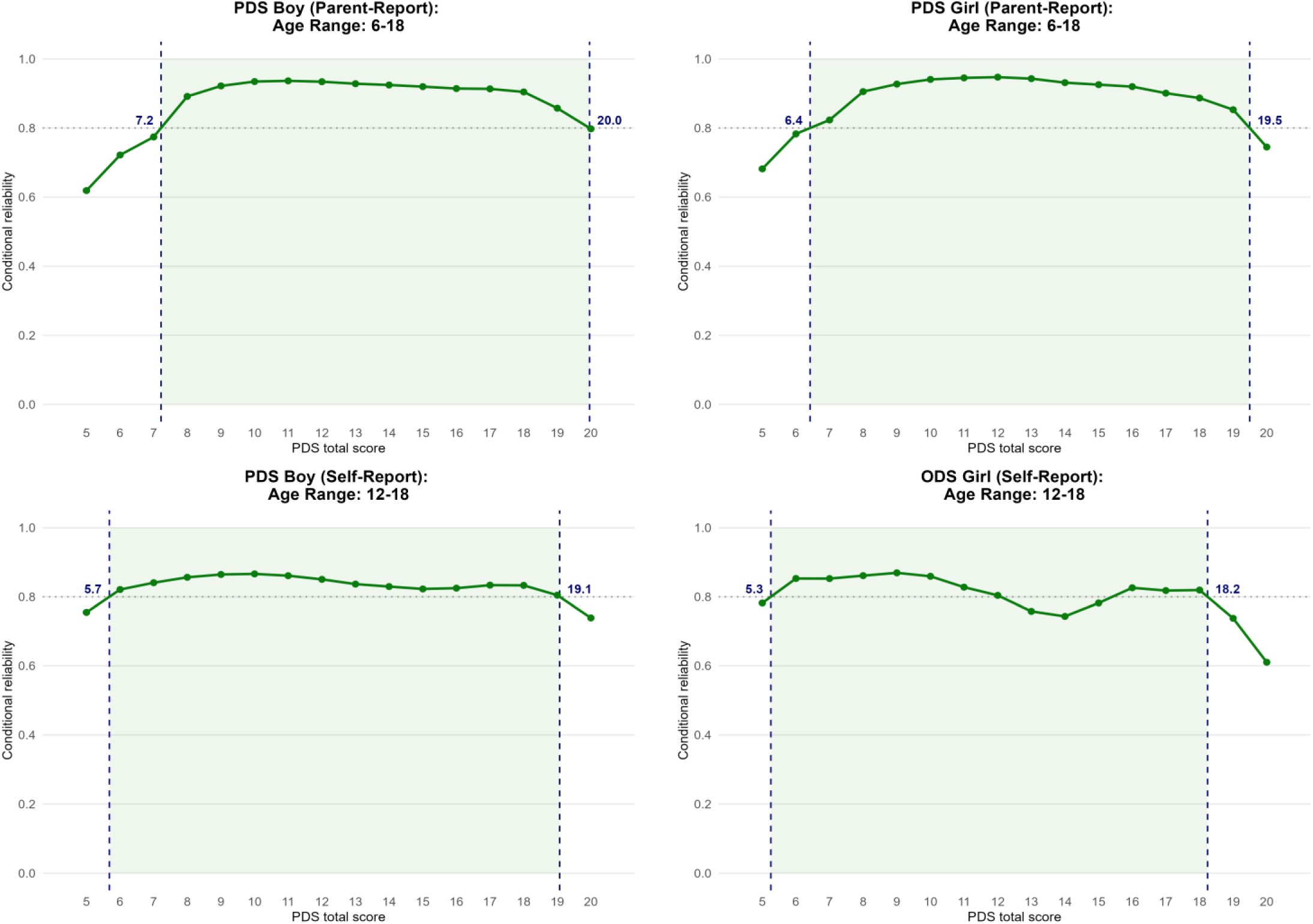
Conditional reliability across the PDS observed score range based on graded response model for all ordinal items with 4-level responses and two-parameter logistic model for the dichotomous menarche item. Expected A Posterior scoring was applied to map the latent trait onto the observed score metric.

For youth self-report PDS, reliability was more attenuated across the score range, consistent with the smaller *α* and *ω* values observed for this version. For boys, reliability exceeded .80 at score 5.7-19.1 (Figure 1C). For girls (Figure 1D), the .80 criterion was met for score 5.3-18.2, though not continuously, with reliability dropping below .80 at score 13-15. This attenuation likely reflects the dichotomous scoring of the menarche item (1 or 4, with no intermediate values), which, on a scale with five items, yields a disproportionate reduction in information density across the pre– to post-menarche transition. Nevertheless, conditional reliability within this attenuated region remained above .75, which is still sufficient for detecting meaningful changes in pubertal development. Overall, the PDS demonstrated adequate precision in measuring the pubertal development construct across an extensive portion of the score range, with reliability attenuated at the parent-report floor and the self-report ceiling.

### Test-Retest Reliability

Test-retest reliability was excellent across informant and child sex (Table 4). For parent-report PDS, both boys (*r*=.95, ICC=.95) and girls (*r*=.96, ICC=.96) showed excellent stability across the approximately 2.5 week retest interval. For youth self-report PDS, reliability was similarly excellent, though modestly lower than parent-report, for boys (*r*=.90, ICC=.89) and girls (*r*=.91, ICC=.90). Across all four groups, Pearson’s *r* and ICC(2,1) values were nearly identical, indicating that PDS total scores did not shift systematically across the retest interval and that the high correlation in relative standing reflected by Pearson’s *r* was matched by comparably high absolute agreement reflected by ICC.

**Table 4.** PDS test-retest reliability across roughly 19 days of retest interval.

| Sex | *n* | Pearson's *r* | 95% CI | ICC(2,1) | 95% CI |
| --- | --- | --- | --- | --- | --- |
| Parent Report |  |  |  |  |  |
| Boy | 88 | .95 | .92-.97 | .95 | .92-.96 |
| Girl | 83 | .96 | .93-.97 | .96 | .94-.97 |
| Youth Self-Report |  |  |  |  |  |
| Boy | 44 | .90 | .82-.94 | .89 | .81-.94 |
| Girl | 45 | .91 | .84-.95 | .90 | .83-.94 |

*Note.* The average retest interval was 18.90 days for parents and 19.39 days for youth. $ICC(2,1)$ : Intraclass Correlation Coefficient based on two-way random effects model for absolute agreement and single unit of analyses.

### Measurement Invariance

Of all item-by-group DIF tests conducted across informant sample (parent-report of children ages 6-11 and 12-18, youth self-report ages 12-18), child sex (male vs. female), and grouping variables (race: White vs. non-White; parental education: bachelor’s degree or higher vs. less than a bachelor’s degree), only four DIF tests reached statistical significance after false discovery rate correction (Table 5).

**Table 5.**
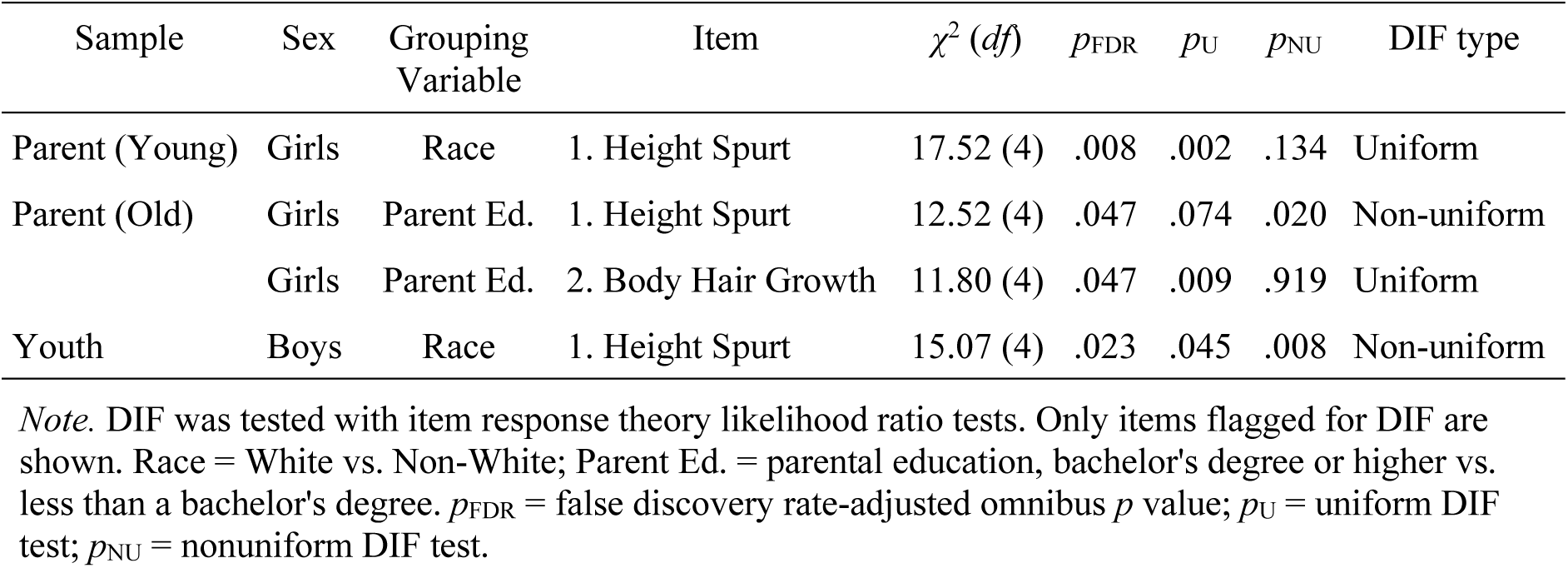
PDS Differential Item Functioning by Race and Parental Education Level.

Among parent-reported girls ages 6-11, Item 1 (height spurt) showed uniform DIF by race (*χ*^2^(4)=17.52, *p*=.008), with parents of non-White girls being expected to endorse a higher height-spurt category than White girls at the same low-to-moderate pubertal development, though the two groups converged at higher trait levels (Figure 2A). Among parent-reported girls ages 12-18, both Item 1 (height spurt; *χ*^2^(4)=12.52, *p*_BH_=.047) and Item 2 (body hair growth; *χ*^2^(4)=11.80, *p*_BH_=.047) showed DIF by parental education: girls whose parents held a bachelor’s degree or higher were expected to endorse slightly higher item scores than girls of less-educated parents at low-to-moderate trait levels, again converging at higher levels of pubertal development (Figure 2B, Figure 2C). Despite these significant item-level effects, DTF analyses indicated that no items produced meaningful distortion at the test level. For the ages 6-11 sample, focal-weighted sDTF and uDTF were –0.13 and 0.13 points of PDS total score, respectively; for the ages 12-18 sample, they were –0.14 and 0.17. Given that the PDS total scores range 5-20, the item-level DIF impact on expected PDS total scores was functionally negligible.

**Figure 2.**
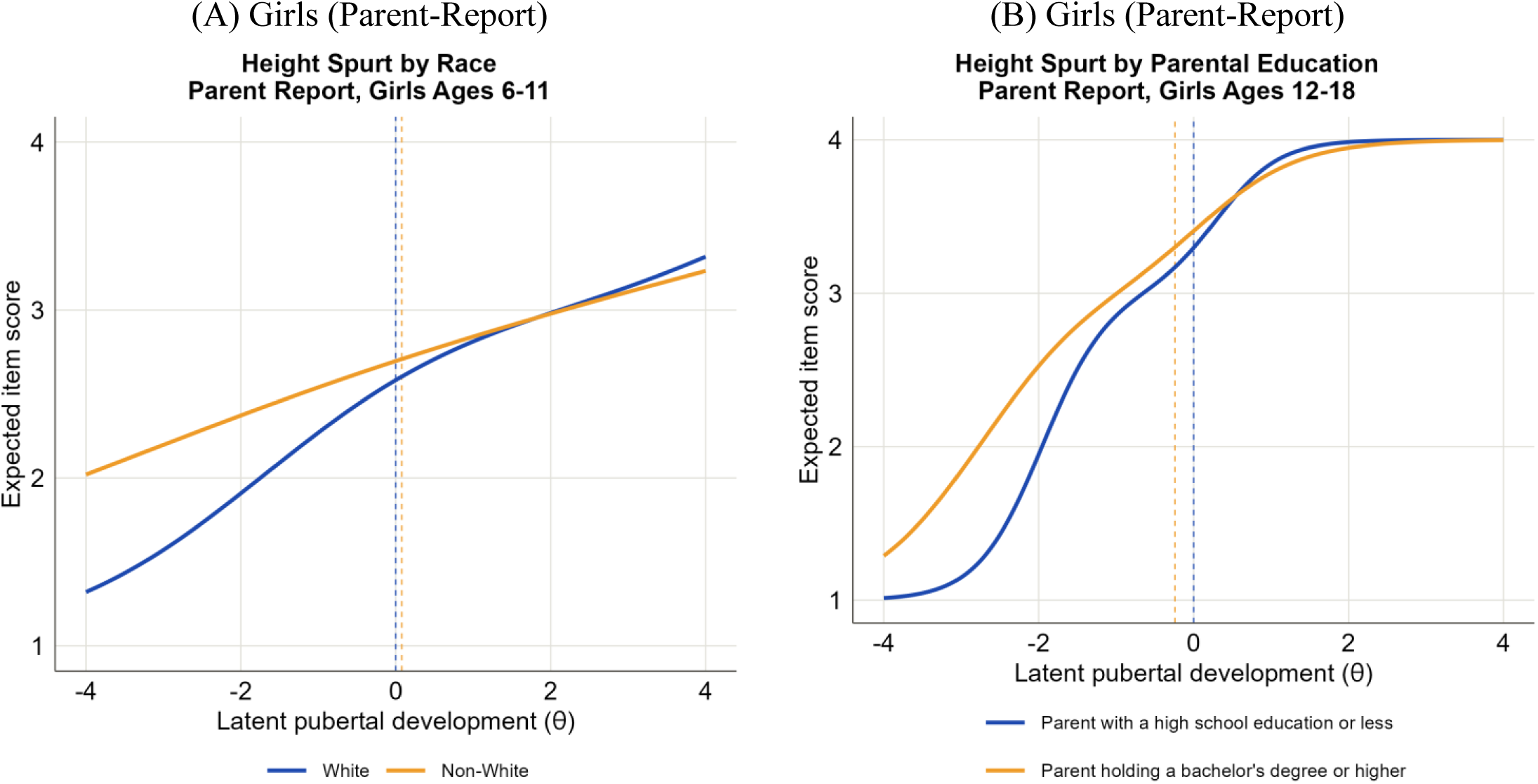
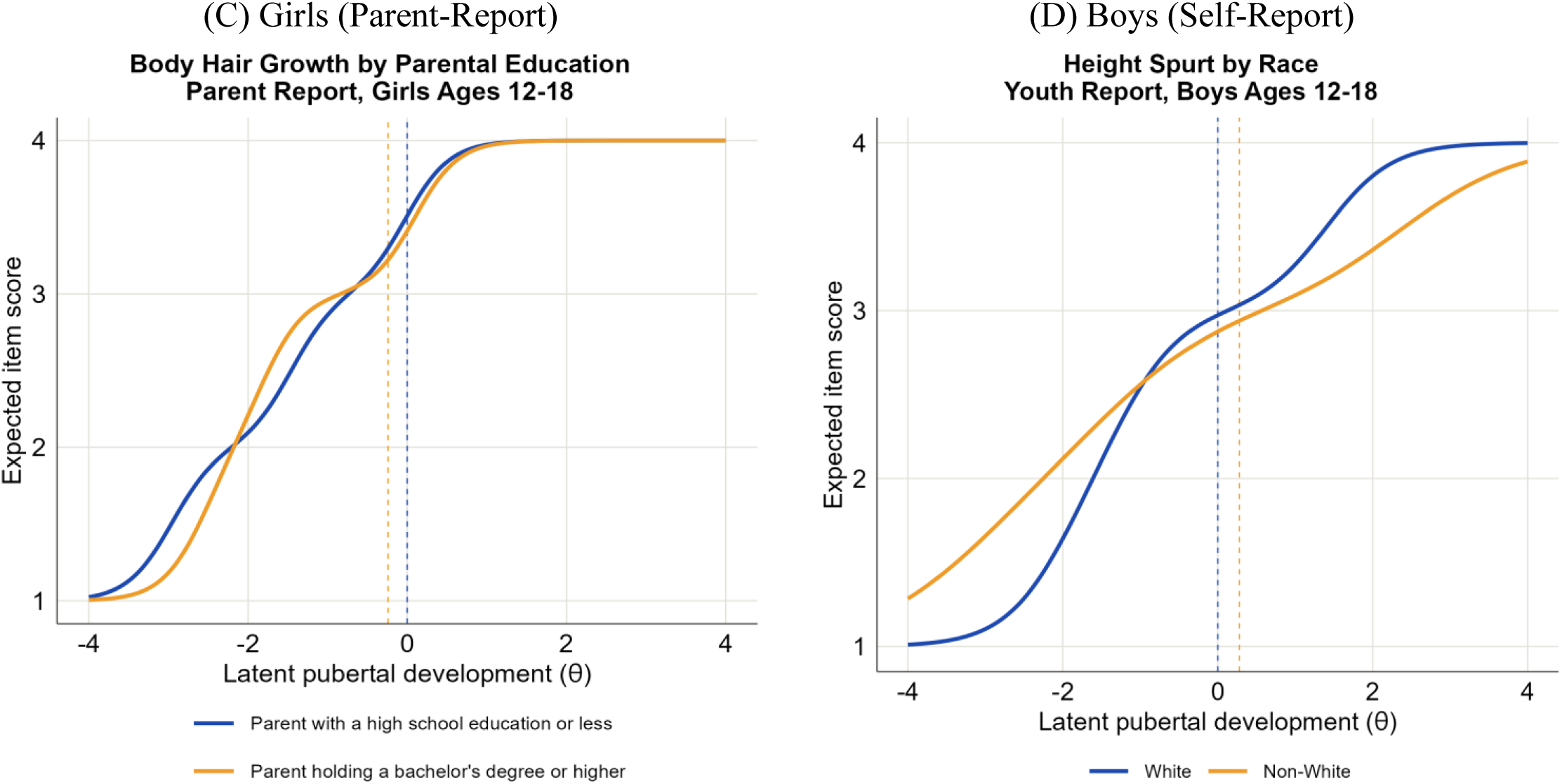
Expected item score curves for Pubertal Development Scale items exhibiting differential item functioning (DIF); dashed vertical lines indicate group mean θ.

Among youth-reported boys ages 12-18, Item 1 (height spurt) showed nonuniform DIF by race (*χ*^2^(4)=15.07, *p*_BH_=.023): non-White boys reported comparatively higher height-spurt scores at low pubertal development, whereas White boys reported higher scores at high pubertal development, reflecting a difference in item discrimination rather than a uniform shift (Figure 2D). Similarly, the DTF analysis suggested minimal test-level difference caused by item DIF with sDTF=.12 and uDTF=.17.

### Cross-Informant Agreement

ICC analyses revealed excellent cross-informant absolute agreement between parents and children in rating the child’s pubertal development (boys: ICC=.88, 95% CI=.85-.90; girls: ICC=.87, 95% CI=.84-.89; see Table 6), meaning that parents and children were highly consistent in their rating, with small variation between informants.

**Table 6.** Cross-informant agreement between parent– and youth-report PDS total scores.

| Cross-Informant Agreement | ICC(2,1) | 95% CI |
| --- | --- | --- |
| Parent-Boy | .88 | .85-.90 |
| Parent-Girl | .87 | .84-.89 |

*Note.* ICC based on two-way random effects model for absolute agreement and single unit of analyses, specified as ICC(2,1).

Bland-Altman plots further supported these high ICC values. For boys (Figure 3A), parents showed a nonsignificant mean difference of –0.07 from their children (paired *t*-test, *p*=0.297), indicating that on average, parents and boys rated pubertal development almost identically. The 95% limits of agreement ranged from –3.07 to 2.93, meaning that for 95% of boy-parent dyads, scores differed by no more than roughly ±3 points. A linear regression of differences on means revealed a small but statistically significant proportional bias (*β*=.061, *p*=.008), indicating that parent-boy difference shifted slightly in the positive direction as PDS mean scores increased. In other words, at lower mean PDS levels, parents tended to rate boys slightly lower than boys rated themselves, whereas at higher levels ratings converged or became slightly higher. However, the magnitude of this effect was minimal, explaining only 1.4% of variance and suggesting limited practical impact. For girls (Figure 3B), parents showed a statistically significant mean difference of –0.13 (paired *t*-test, *p*<.05), but this small difference is clinically negligible given that the PDS total scores range 5-20. The 95% limits of agreement were from –2.59 to 2.34, which is a narrower spread than observed for boys. No evidence of proportional bias was found (*β*=.012, *p*=.606), suggesting that parent-girl differences remained consistent across the PDS score range.

**Figure 3.**
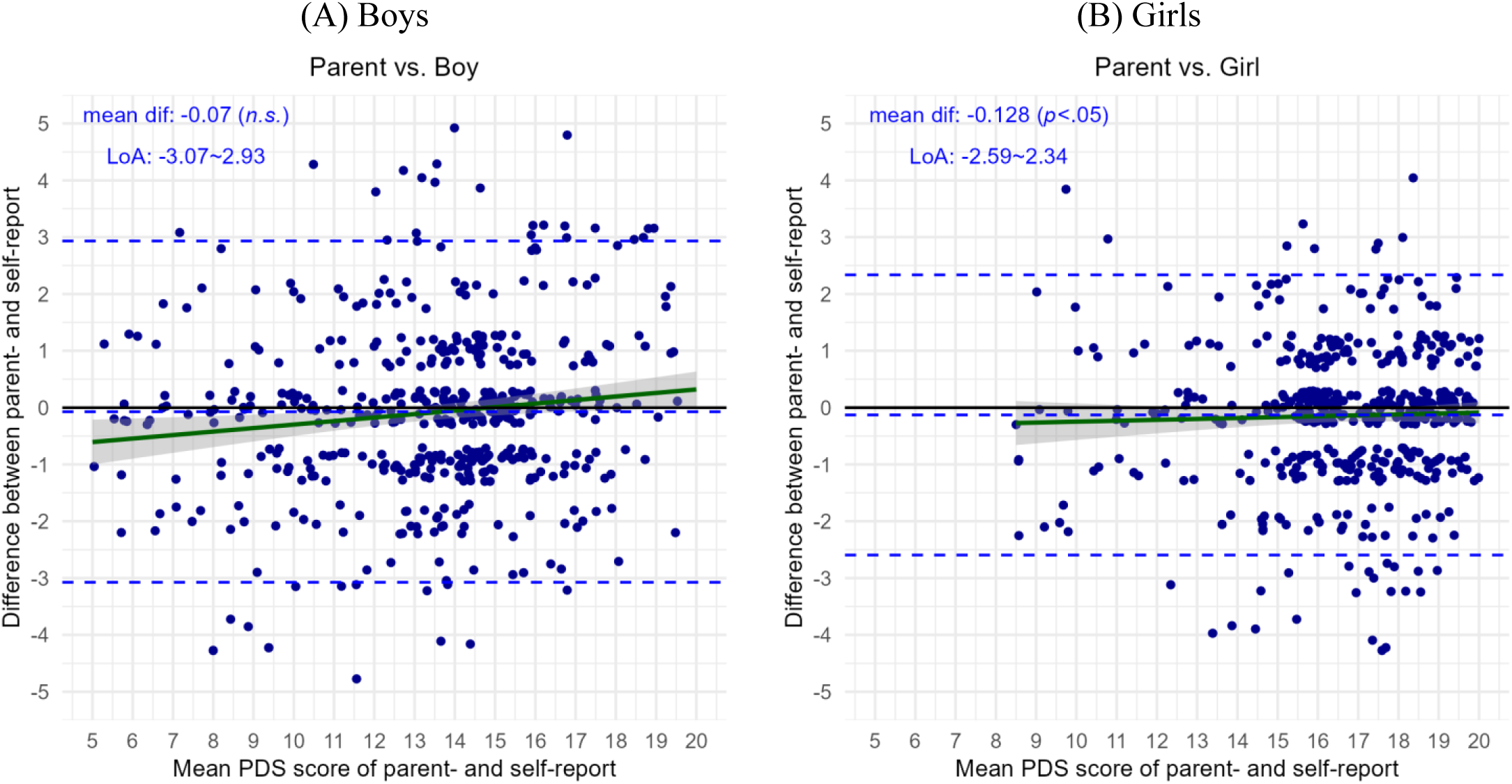
Bland-Altman plots depicting parent-youth agreement on PDS total scores, with mean difference, limits of agreement (LoA), and linear regression line of differences on means displayed. Panel A: Parent vs. Boy. Panel B: Parent vs. Girl.

Together, the ICC analyses and Bland-Altman plots indicated that for children aged 12-18, parent-report and self-report PDS scores demonstrate strong agreement with acceptable ranges of difference.

### Parent-Report Norms

Based on GAMLSS percentile curves (see Figure 4), parent-report PDS scores increased nonlinearly with age and exhibited sex-specific variance patterns. Among boys (Figure 4A), the median score increased from 7 at age 6 to 18 at age 18, with the steepest growth occurring between ages 11-15. The 5^th^-95^th^ percentile range was narrow during prepuberty (5-8 points at ages 6-10), expanded during peak development (7-10 points at ages 11-15), and remained wide through adolescence (7-8 points at ages 16-18). Even at age 18, 90% of boys still scored between 13 and 20, suggesting substantial heterogeneity in boys’ pubertal timing at late adolescence.

**Figure 4.**
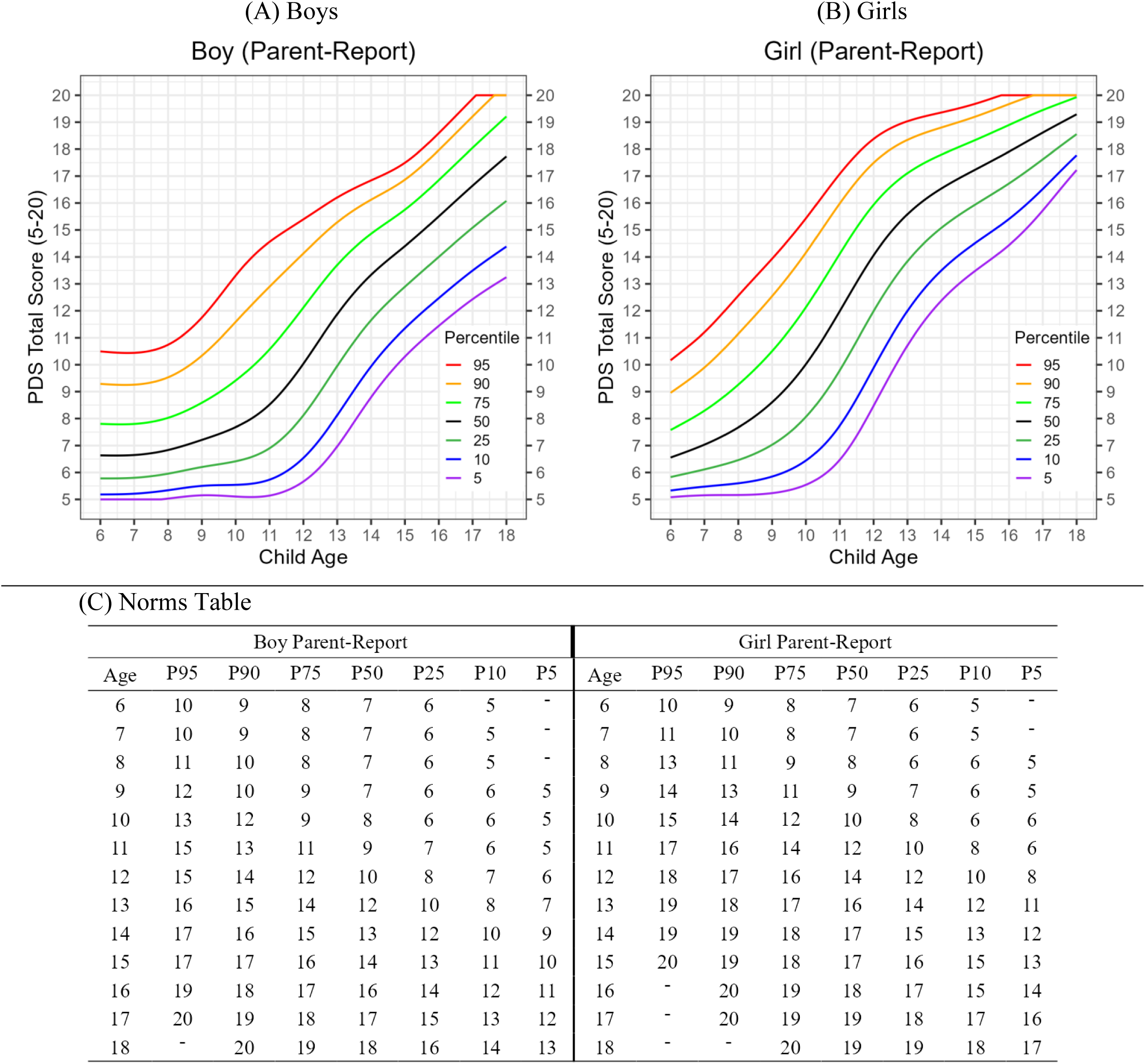
Parent-report PDS norms. GAMLSS-estimated percentile curves (5^th^-95^th^) for parent-report PDS total scores (range: 5-20) across ages 6-18 years, shown for (A) boys and (B) girls, with (C) the reference percentile table.

In contrast, girls exhibited earlier pubertal onset and faster progression (Figure 4B). The median score rose from 7 at age 6 to 19 at age 18, and the peak growth occurred between ages 9-13, showing a two-year advancement compared to boys that is consistent with established biological evidence (Brix et al., 2019). The 5^th^-95^th^ percentile began widening as early as age 7, reached maximum dispersion of 11 points around age 11, then gradually narrowed after age 13. By age 18, 90% of girls scored between 17 and 20, reflecting more uniform pubertal completion among girls. The norms table in Figure 4C presents key percentile estimates rounded to integers of each age-sex combination. Parent-reported percentile curves and reference table rescaled to the PDS average score on a 1-4 unit can be found in Supplemental Figure S2.

### Youth Self-Report Norms

Youth self-report PDS scores demonstrated developmental patterns similar to parent-report across the overlapping age range of 12-18 years (see Figure 5). For boys (Figure 5A), the median score rose from 11 at age 12 to 17 at age 18. Dispersion, reflected by the 5^th^-95^th^ percentile gap, was widest at age 12 (10 points) and narrowed to 6 points by age 18. For girls (Figure 5B), the median score rose from 15 at age 12 to 19 at age 18, and the dispersion decreased from 8 points to 3 points across this age range. Girls again demonstrated greater convergence in pubertal completion than boys by age 18, regardless of informant type. Key percentile estimates are presented in the norms table in Figure 5C. Rescaled self-reported norm curves and table can be found in Supplemental Figure S3.

**Figure 5.**
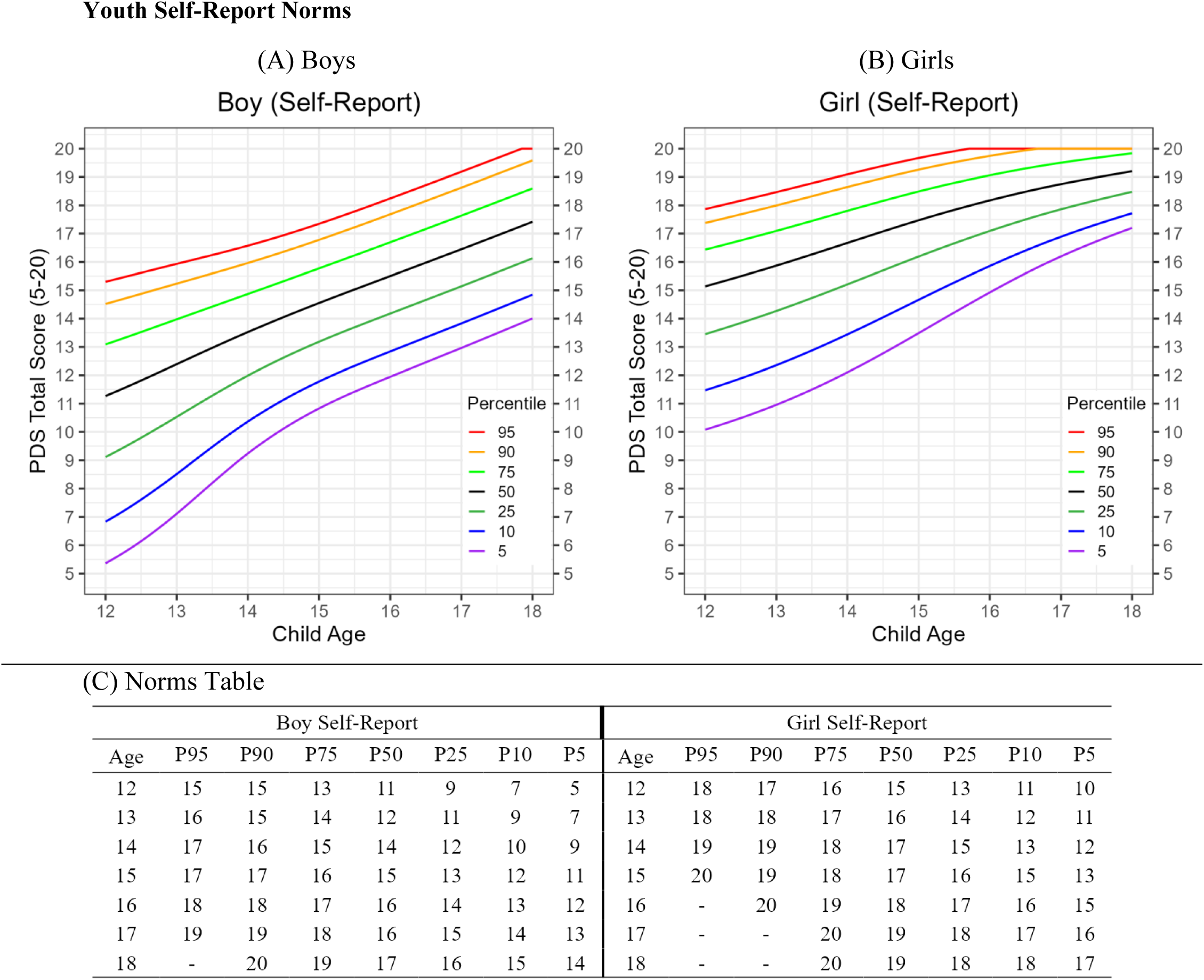
Youth self-report PDS norms. GAMLSS-estimated percentile curves (5^th^-95^th^) for self-report PDS total scores (range: 5-20) across ages 12-18 years, shown for (A) boys and (B) girls, with (C) the reference percentile table.

### Pubertal Timing and Clinical Symptoms

After accounting for child age and sex, pubertal timing, represented as age– and sex-normed PDS percentile, was independently associated with parent-reported clinical symptoms (Table 7). With all symptom scores and PDS percentiles standardized as z-scores within the full sample, earlier pubertal timing relative to peers predicted higher parent-reported depression (*β*=.26, *p*<.01), mania (*β*=.46, *p*<.001), and sleep disturbance (*β*=.40, *p*<.001), but not hyperactivity (*β*=.15, *ns*) or inattention (*β*=.03, *ns*). Each of these significant associations was accompanied by a significant negative age-by-pubertal-timing interaction (*β*s=-.02 to –.03, *p*s<.01), indicating that the association between earlier pubertal timing and elevated clinical symptoms was strongest among younger children and attenuated with increasing age. A significant female-by-pubertal-timing interaction was additionally observed for parent-reported mania (*β*=-.10, p<.05), suggesting that the positive association between earlier pubertal timing and elevated mania symptoms was more pronounced among boys. Variance explained by the parent-report models ranged from 1.1% (inattention) to 7.4% (hyperactivity), with the hyperactivity prediction driven primarily by strong age and sex main effects.

**Table 7.** Regression Estimates Predicting Clinical Symptoms from Child Age, Sex, and Pubertal Timing.

| Outcome | Age | PT | Female | Age × PT | Female × PT | $R^2$ |
| --- | --- | --- | --- | --- | --- | --- |
| **Parent Report** |  |  |  |  |  |  |
| Depression | 0.030*** | 0.255** | 0.071 | -0.018** | -0.026 | 1.7% |
| Mania | -0.016* | 0.461*** | -0.088 | -0.034*** | -0.095* | 2.1% |
| Hyperactivity | -0.067*** | 0.145 | -0.246*** | -0.017* | 0.013 | 7.4% |
| Inattention | -0.008 | 0.028 | -0.189*** | -0.005 | 0.041 | 1.1% |
| Sleep | 0.063*** | 0.398*** | 0.016 | -0.030*** | -0.026 | 5.8% |
| **Youth Report** |  |  |  |  |  |  |
| Depression | -0.020 | 0.503 | 0.246** | -0.029 | -0.018 | 2.4% |
| Mania | -0.007 | 0.463 | 0.063 | -0.031 | -0.060 | 0.5% |
| Hyperactivity | -0.063* | 0.376 | -0.059 | -0.031 | 0.081 | 1.7% |
| Inattention | -0.056* | 0.157 | -0.008 | -0.010 | 0.060 | 1.1% |
| Sleep | 0.043 | 0.488 | 0.158 | -0.032 | 0.000 | 1.6% |

*Note.* Clinical symptom scores and PDS percentiles were standardized as a $z$ -score ( $M=0$ , $SD=1$ ) to facilitate comparison of regression estimates across outcomes. Rows are grouped by informant. PT=pubertal timing; $R^2$ = proportion of variance explained. All $p$ -values were adjusted for false discovery rate. Significance level: \* $p<.05$ , \*\* $p<.01$ , \*\*\* $p<.001$

In the youth self-report sample, no significant associations between pubertal timing and the five clinical outcomes were detected after false discovery rate correction, though point estimates were of comparable magnitude (*β*=.16-.50). This lack of significance is more likely to be attributable to the distinct reporting perspective inherent in youth self-report measures rather than differences in sample size: the smaller youth sample (*N*=1006) had 95% power to detect correlations of .11, versus .08 for the larger parent sample (*N*=2251), based on a GPower calculation (Erdfelder et al., 1996). Female sex was associated with higher self-reported depression (*β*=.25, *p*<.01), and younger age predicted higher hyperactivity (*β*=-.06, *p*<.05) and inattention (*β*=-.06, *p*<.05). *R*^2^ values for the youth-report models were modest, ranging from 0.5% (mania) to 2.4% (depression).

Together, these findings suggest that norm-referenced pubertal timing carries predictive values about children’s mental and behavioral health beyond chronological age and sex, an association most clearly detected through parent report and concentrated in depression, mania, and sleep disturbance.

## Discussion

In this study, we examined the psychometrics of the Pubertal Development Scale and established its continuous norms in a nationally representative sample of U.S. children. We validated the scale with modern measurement methods in a broad age range of children ages 6-18 for parent-report and children ages 12-18 for self-report. The age– and sex-specific norms generated through the GAMLSS approach for both informants, in the format of percentile curves and reference tables, provide precise pubertal timing indices that enhance the scale’s research and clinical utility.

The factor analyses confirmed that the PDS captures a single, coherent pubertal development construct across informants and sexes, supporting the field’s standard practice of summing or averaging items into a single score. When floor effects among prepubertal children were accounted for by restricting parent-report models to an age-appropriate window, the one-factor structure strongly fit the PDS data, which extends prior validation work to a more representative age range. Acceptable-to-good internal consistency was demonstrated for both informants regardless of child sex, with Cronbach’s *α* ranging .77-.89 and McDonald’s *ω* ranging .78-.90. Regarding measurement precision examined through the IRT framework, conditional reliability exceeded .80 across most of the score continuum, spanning roughly 13 of the 15 possible total-score points. Reliability attenuations were concentrated at the parent-report floor and the self-report ceiling, reflecting limits in item-response variability among children with minimal or near-complete pubertal development. Test-retest reliability was excellent across informants and child sex (*r*=.90-.96; ICC=.89-.96), and the close correspondence between Pearson’s *r* and ICC indicates that PDS scores remained stable in both relative standing and absolute level over the roughly 19-day retest interval. Taken together, the PDS yields reliable and accurate scores within a single administration and across repeated occasions.

Measurement invariance testing showed that the PDS performs comparably across demographic subgroups. Although a small number of items exhibited statistically significant DIF by race or parental education, these effects did not produce meaningful distortion at the test-score level, indicating that total PDS scores can be interpreted similarly regardless of a child’s sociodemographic background. These psychometric findings support the PDS as a valid and reliable measure of pubertal development suitable for large-scale research and clinical use, while also clarifying the age ranges and score regions where its precision is strongest.

We also verified that parent– and self-report PDS had excellent cross-informant agreement at ages 12-18 (ICC: .87-.88). Based on this finding, researchers and clinicians could select informants based on their clinical questions and contextual needs. Parent-report PDS, covering a broader age range of 6-18, is preferable when the population of interest includes younger children for whom the self-assessment is less accurate (Peng et al., 2018) and normative references are unavailable. Parent-report is also the most viable option for detecting precocious puberty, defined as pubertal onset before age 8 in girls and age 9 in boys (Bradley et al., 2020), since children at this age might lack the bodily awareness and vocabulary to report on pubertal changes. Self-report PDS works well with adolescents capable of reliable self-assessment, particularly in contexts that prioritize patient privacy or direct clinician-adolescent interaction. The PDS, with parent– and self-report formats, is a flexible measure that can be adapted to diverse research and clinical settings.

The national norms for the PDS offer important benchmarks to advance the interpretation of PDS scores and to aid in the identification of atypical pubertal timing. Pubertal development exhibited sex differences, age-dependent changes, and inter-individual heterogeneity, reflected in the nonlinear trajectories and varying spread of PDS scores across age within each sex. The GAMLSS norms effectively modeled these sources of variations and estimated 5^th^-95^th^ percentiles to contextualize the PDS. Researchers and clinicians can refer to the median curve (50^th^ percentile) as the expected pubertal trajectory and use the 90^th^ and 10^th^ percentiles to signal early or delayed puberty. The GAMLSS norms positioned the PDS within a dynamic interpretative framework that examines more than a score’s face value but also its developmental meaning given a child’s age and sex.

In addition to establishing the PDS as a psychometrically sound measure, this study demonstrates that norm-referenced pubertal timing carries important clinical predictive value. In our sample, children with earlier pubertal development showed elevated parent-reported depressive and manic symptoms and sleep disturbance, independent of their chronological age and sex. With the examination of age-by-pubertal-timing interactions, we also found that the associations between early pubertal timing and clinical symptoms were strongest in younger children and weakened with age. Our findings are consistent with a broader literature linking earlier pubertal timing to depression risk (Benson et al., 2026) and, more tentatively, to sleep disruption (Kansagra, 2020). The mania association we found is comparatively novel, with existing evidence linking pubertal hormones and manic-spectrum symptoms largely mechanistic and untested (Colic et al., 2025). In the youth self-report sample, we did not detect significance of these correlations despite comparable effect sizes, which likely reflects differences in informant perspective about emotional and behavior symptoms. Clinically, these findings suggest that the age– and sex-normed PDS percentile offers a strong index for flagging children who may be at elevated risk for internalizing and externalizing symptoms, and could inform identification and support for youths experiencing off-time puberty.

Beyond its research applications, the PDS norms fill a practical gap in outpatient mental health settings: Tanner staging already has population-referenced percentile norms embedded in standard growth charts, but mental health workers are rarely positioned to conduct the physical examination it requires. The percentile curves and reference tables reported here provide an analogous benchmark: a psychologist, pediatrician, or behavioral health clinician can administer the parent– or self-report PDS in a standard visit and determine whether a child’s pubertal development falls within an expected range for their age and sex or warrants referral for further evaluations like Tanner staging or endocrine assessment. The PDS percentile is not a standalone diagnostic tool, but a low-burden adjunct to standard symptom measures that can prompt closer monitoring or a referral conversation when early or atypical pubertal change coincides with emerging mood dysregulation (Youngstrom, 2026).

### Limitations

We acknowledge several limitations in this study. First, the PDS psychometrics and norms were derived from population survey data without cross-validation against the clinician-rated Tanner staging. Therefore, we recommend using the PDS scale along with normative benchmarks as a broad estimation of pubertal development in research contexts or as a pre-screener to warrant further assessment in pediatric settings. Second, although the 6-18 age range of the current study offers a broader coverage than prior studies, it may not fully capture the complete pubertal development period for all youths. Notably, boys continued to show substantial heterogeneity in PDS scores at age 18, suggesting that pubertal completion could extend beyond the upper age limit of the current norms. Future studies should consider expanding the age range to better represent late-developing adolescents. Third, although our survey data is by far the most nationally representative, its demographic characteristics still deviated from the 2023 ACS estimates, particularly in the child racial composition and parental education level. It is worth noting that households with children, the focal population of our study, may differ demographically from the general U.S. population represented by ACS estimates. Nevertheless, our measurement invariance analysis based on Differential Item and Test Functioning confirmed that the PDS functions similarly across the racial (White vs. non-White) and education level (bachelor’s degree or higher vs. less than a bachelor’s degree) grouping variables, supporting its adoption in subpopulations underrepresented in our data.

### Conclusion

In conclusion, this study establishes national psychometrics and continuous norms for the PDS, filling a gap in the standardization of this puberty scale. By validating both parent– and self-report versions in a nationally representative sample and generating sex-specific normative benchmarks, we provide researchers and clinicians an empirical foundation for interpreting PDS scores. Our findings support the PDS’s continued use as an accessible, flexible tool for characterizing pubertal development and identifying youths with atypical pubertal timing.

## Data Availability

Code for replication is available at https://github.com/YinuoLiu0708/5040.11-PDS-psychometrics-norms.git. Data are available upon request from the corresponding author.

## Author Note

Data Availability Statement: Code for replication is available at https://github.com/YinuoLiu0708/5040.11-PDS-psychometrics-norms.git. Data are available upon request from the corresponding author.

Ethics Approval: The IRB of Nationwide Children’s Hospital provided an NHSR determination for analyses of deidentified survey data.

Conflict of Interest: Eric A. Youngstrom is the co-founder and Executive Director of Helping Give Away Psychological Science, a 501c3; he has consulted about psychological assessment with Signant Health and received royalties from the American Psychological Association and Guilford Press, and he holds equity in Joe Startup Technologies and held equity in Autism Intervention Measures.

Author roles were classified using the Contributor Role Taxonomy (CRediT; https://credit.niso.org/) as follows: *Yinuo Liu*: conceptualization, data curation, methodology, investigation, formal analysis, and writing – original draft. *Andrea E. Bonny*: review & editing. *Guillermo Perez Algorta*: methodology and review & editing. *Eric A. Youngstrom*: supervision, methodology, resources, validation, and review & editing

