## supplemental material for "National Norms and Psychometrics for the Pubertal Development Scale"

### Author Note

Yinuo Liu https://orcid.org/0009-0007-2606-575X

Andrea E. Bonny https://orcid.org/0000-0001-5874-1942

Eric A. Youngstrom https://orcid.org/0000-0003-2251-6860

**Table S1**
Demographic characteristics of the Retest sample

| | Parent Retest ($N$ = 171) | Youth Retest ($N$ = 90) |
|---|---|---|
| N | 171 | 90 |
| Child age | | |
| M (SD) | 11.53 (3.24) | 14.37 (1.67) |
| 6-7 years | 15.8% | |
| 8-9 years | 17.0% | |
| 10-11 years | 12.9% | |
| 12-13 years | 24.0% | 36.7% |
| 14-15 years | 18.1% | 33.3% |
| 16-18 years | 12.3% | 30.0% |
| Child sex | | |
| Female | 48.5% | 51.1% |
| Male | 51.5% | 48.9% |
| Race | | |
| White | 77.8% | 70.0% |
| Black or African American | 7.0% | 12.2% |
| Asian | 3.5% | 1.1% |
| American Indian or Alaska Native | 0.6% | 3.3% |
| Native Hawaiian or Pacific Islander | 0.6% | 1.1% |
| Some other race | 4.1% | 2.2% |
| Multiple races selected | 5.3% | 10.0% |
| Ethnicity | | |
| Hispanic, Latino, or Spanish Origin | 14.4% | 17.1% |
| Not Hispanic, Latino, or Spanish Origin | 85.6% | 82.9% |
| Parent/guardian education | | |
| High school graduate or higher | 97.1% | 100.0% |
| Bachelor's degree or higher | 40.9% | 53.3% |
| Median Household Income | $70,000 - $79,999 | $70,000 - $79,999 |

**Table S2**
Global fitness comparison for the PDS GAMLSS models

| | BIC | |
|---|---|---|
| Distribution | Boys | Girls |
| *Parent report* | | |
| BCCG | **5303.03** | **4705.16** |
| t-family | 5398.04 | 4807.99 |
| Normal | 5448.03 | 4824.44 |
| Beta-Binomial | 5423.54 | 4725.19 |
| Binomial | 5496.52 | 4875.83 |
| *Youth self-report* | | |
| BCCG | 2297.95 | 2032.91 |
| t-family | 2315.80 | 2092.53 |
| Normal | 2312.35 | 2098.96 |
| Beta-Binomial | **2289.98** | **2013.81** |
| Binomial | 2306.97 | 2044.65 |

*Note.* Bayesian Information Criterion (BIC) for each candidate score distribution, fit separately by informant and sex. Lower values indicate better fit; the lowest BIC in each column within an informant block is shown in bold. BCCG = Box-Cox Cole and Green distribution.

(A) PDS Boy (Parent-Report)

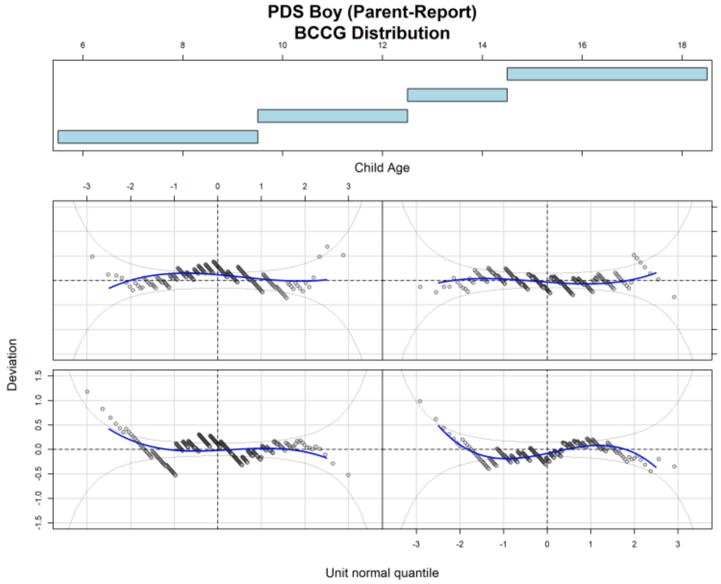


(B) PDS Girl (Parent-Report)

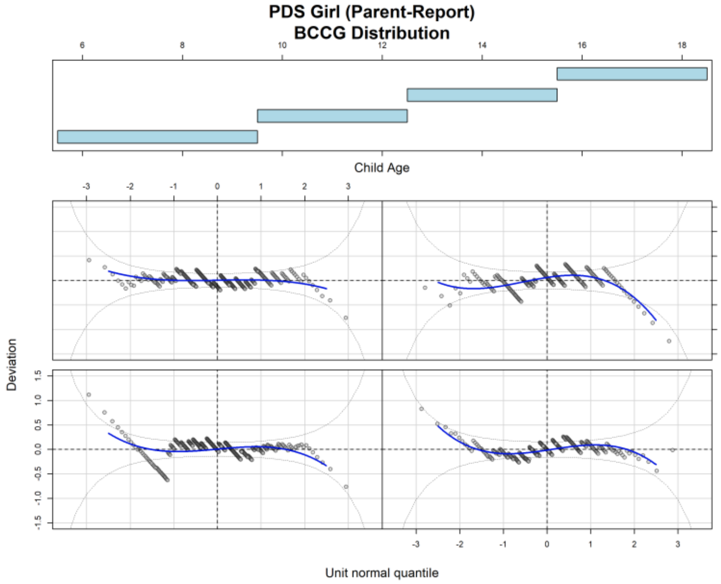


(C) PDS Boy (Self-Report)

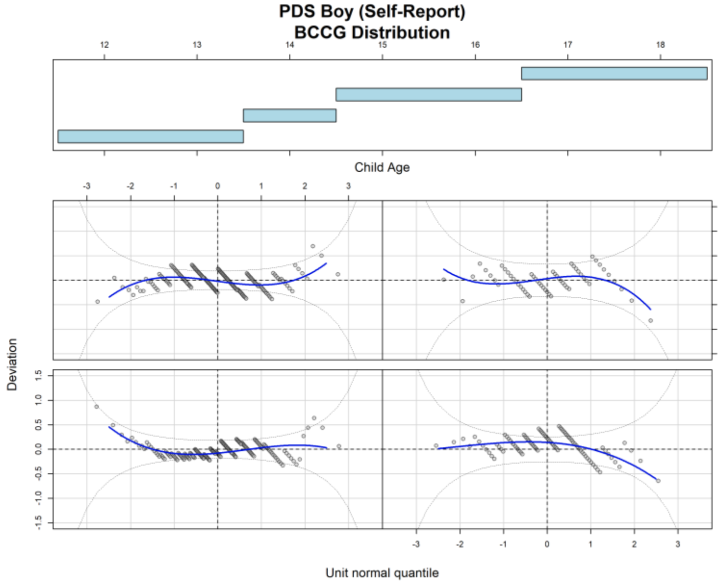


(D) PDS Girl (Self-Report)

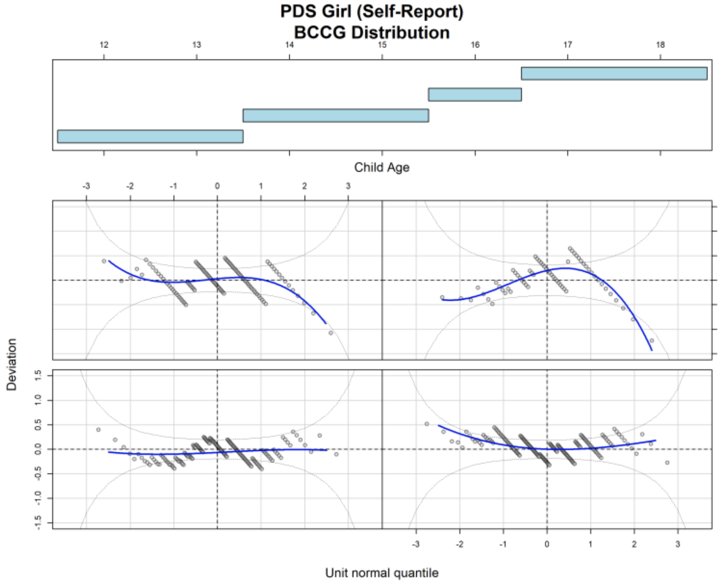


**Figure S1**

Worm plots of normalized quantile residuals for PDS BCCG models, stratified by age window. The smoothed curve within each panel is the primary fit diagnostic, and point banding reflects the discrete integer structure of PDS scores.

(A) PDS Boy (Parent-Report)

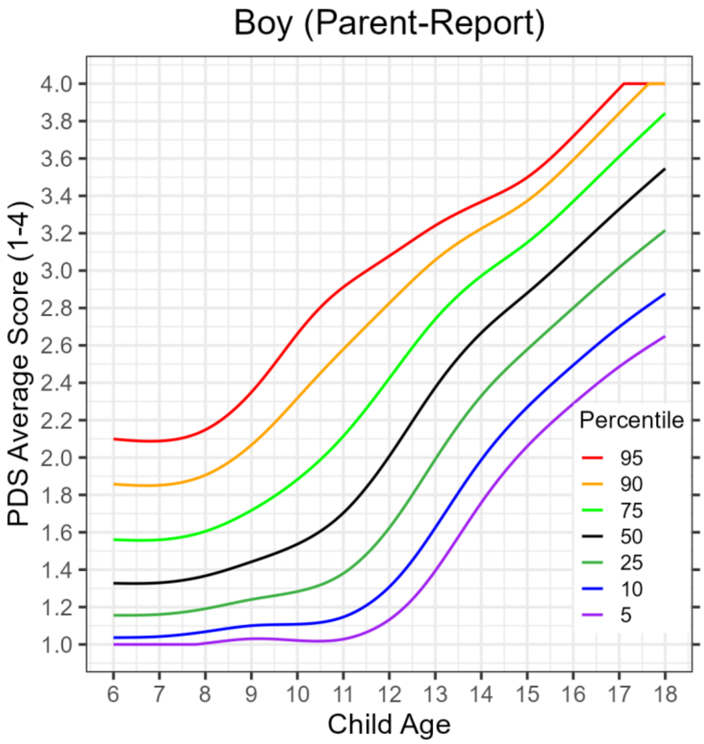


(B) PDS Girl (Parent-Report)

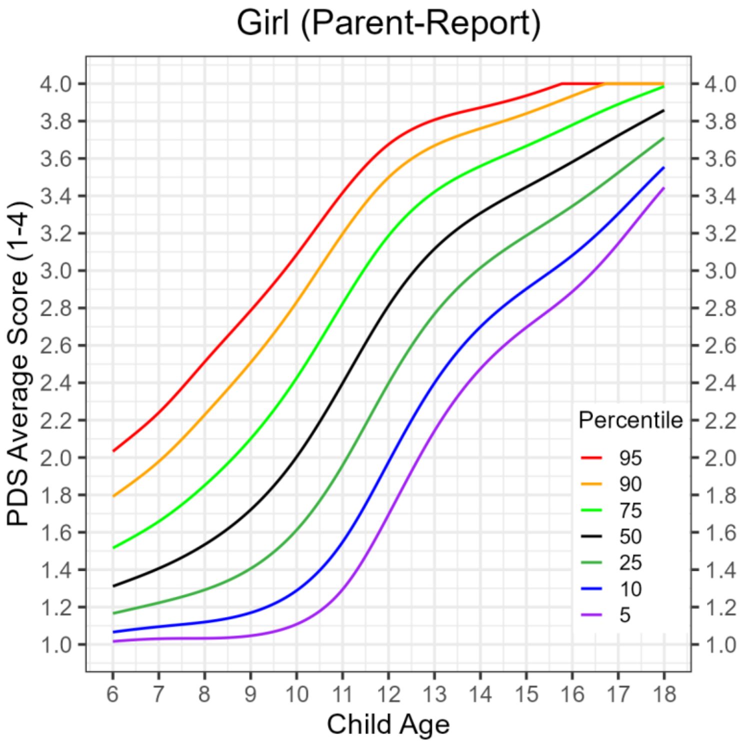


(C) Norms Table

| Boy Parent-Report | | | | | | | | Girl Parent-Report | | | | | | | |
|---|---|---|---|---|---|---|---|---|---|---|---|---|---|---|---|
| Age | P95 | P90 | P75 | P50 | P25 | P10 | P5 | Age | P95 | P90 | P75 | P50 | P25 | P10 | P5 |
| 6 | 2.00 | 1.80 | 1.60 | 1.40 | 1.20 | 1.00 | - | 6 | 2.00 | 1.80 | 1.60 | 1.40 | 1.20 | 1.00 | - |
| 7 | 2.00 | 1.80 | 1.60 | 1.40 | 1.20 | 1.00 | - | 7 | 2.20 | 2.00 | 1.60 | 1.40 | 1.20 | 1.00 | - |
| 8 | 2.20 | 2.00 | 1.60 | 1.40 | 1.20 | 1.00 | - | 8 | 2.60 | 2.20 | 1.80 | 1.60 | 1.20 | 1.20 | 1.00 |
| 9 | 2.40 | 2.00 | 1.80 | 1.40 | 1.20 | 1.20 | 1.00 | 9 | 2.80 | 2.60 | 2.20 | 1.80 | 1.40 | 1.20 | 1.00 |
| 10 | 2.60 | 2.40 | 1.80 | 1.60 | 1.20 | 1.20 | 1.00 | 10 | 3.00 | 2.80 | 2.40 | 2.00 | 1.60 | 1.20 | 1.20 |
| 11 | 3.00 | 2.60 | 2.20 | 1.80 | 1.40 | 1.20 | 1.00 | 11 | 3.40 | 3.20 | 2.80 | 2.40 | 2.00 | 1.60 | 1.20 |
| 12 | 3.00 | 2.80 | 2.40 | 2.00 | 1.60 | 1.40 | 1.20 | 12 | 3.60 | 3.40 | 3.20 | 2.80 | 2.40 | 2.00 | 1.60 |
| 13 | 3.20 | 3.00 | 2.80 | 2.40 | 2.00 | 1.60 | 1.40 | 13 | 3.80 | 3.60 | 3.40 | 3.20 | 2.80 | 2.40 | 2.20 |
| 14 | 3.40 | 3.20 | 3.00 | 2.60 | 2.40 | 2.00 | 1.80 | 14 | 3.80 | 3.80 | 3.60 | 3.40 | 3.00 | 2.60 | 2.40 |
| 15 | 3.40 | 3.40 | 3.20 | 2.80 | 2.60 | 2.20 | 2.00 | 15 | 4.00 | 3.80 | 3.60 | 3.40 | 3.20 | 3.00 | 2.60 |
| 16 | 3.80 | 3.60 | 3.40 | 3.20 | 2.80 | 2.40 | 2.20 | 16 | - | 4.00 | 3.80 | 3.60 | 3.40 | 3.00 | 2.80 |
| 17 | 4.00 | 3.80 | 3.60 | 3.40 | 3.00 | 2.60 | 2.40 | 17 | - | 4.00 | 3.80 | 3.80 | 3.60 | 3.40 | 3.20 |
| 18 | - | 4.00 | 3.80 | 3.60 | 3.20 | 2.80 | 2.60 | 18 | - | - | 4.00 | 3.80 | 3.80 | 3.60 | 3.40 |

**Figure S2**

PDS Parent-report PDS norms rescaled to 1-4 metric. GAMLSS-estimated percentile curves ($5^{th}$-$95^{th}$) for parent-report PDS average scores (range: 1-4) across ages 6-18 years, shown for (A) boys and (B) girls, with (C) the reference percentile table.

(A) PDS Boy (Self-Report)

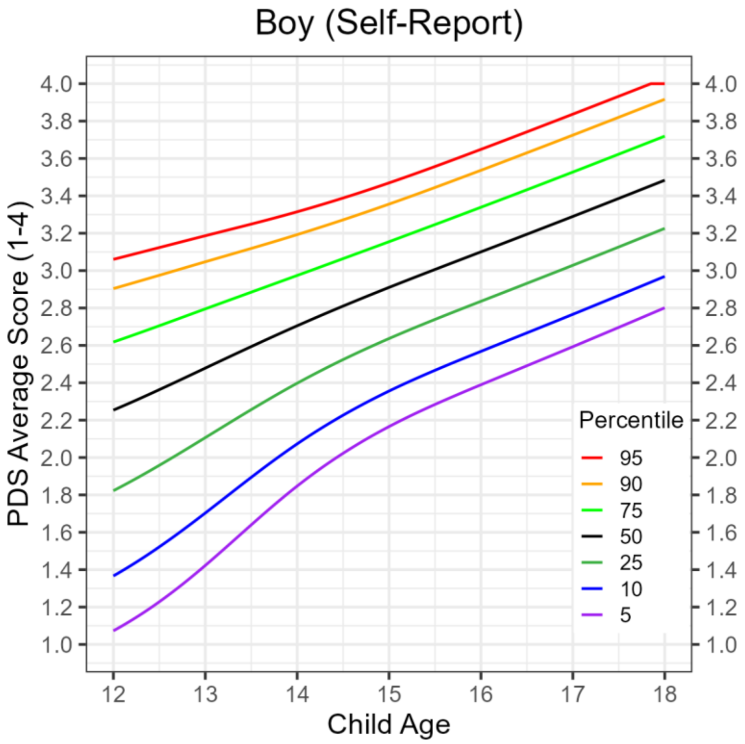


(B) PDS Girl (Self-Report)

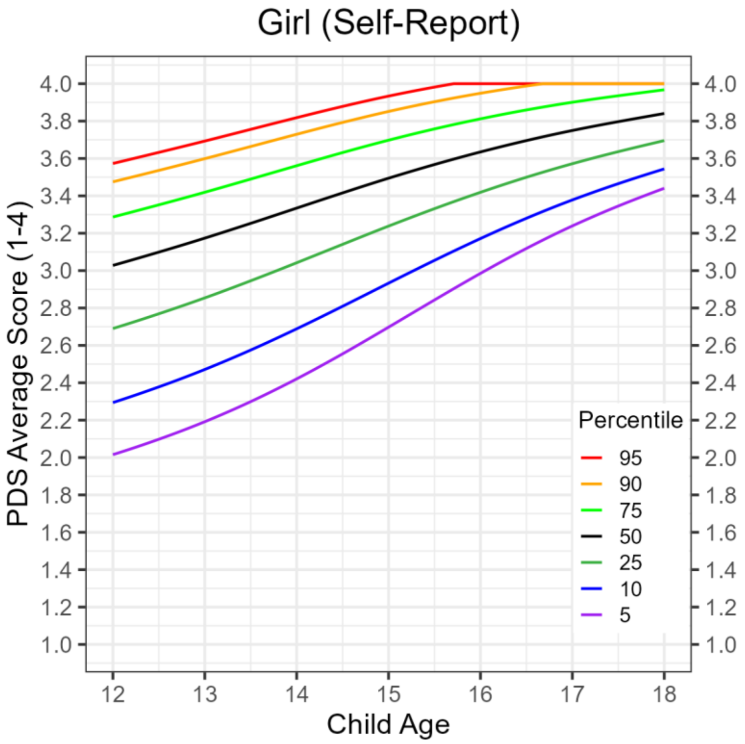


(C) Norms Table

| Boy Self-Report | | | | | | | | Girl Self-Report | | | | | | | |
|---|---|---|---|---|---|---|---|---|---|---|---|---|---|---|---|
| Age | P95 | P90 | P75 | P50 | P25 | P10 | P5 | Age | P95 | P90 | P75 | P50 | P25 | P10 | P5 |
| 12 | 3.00 | 3.00 | 2.60 | 2.20 | 1.80 | 1.40 | 1.00 | 12 | 3.60 | 3.40 | 3.20 | 3.00 | 2.60 | 2.20 | 2.00 |
| 13 | 3.20 | 3.00 | 2.80 | 2.40 | 2.20 | 1.80 | 1.40 | 13 | 3.60 | 3.60 | 3.40 | 3.20 | 2.80 | 2.40 | 2.20 |
| 14 | 3.40 | 3.20 | 3.00 | 2.80 | 2.40 | 2.00 | 1.80 | 14 | 3.80 | 3.80 | 3.60 | 3.40 | 3.00 | 2.60 | 2.40 |
| 15 | 3.40 | 3.40 | 3.20 | 3.00 | 2.60 | 2.40 | 2.20 | 15 | 4.00 | 3.80 | 3.60 | 3.40 | 3.20 | 3.00 | 2.60 |
| 16 | 3.60 | 3.60 | 3.40 | 3.20 | 2.80 | 2.60 | 2.40 | 16 | - | 4.00 | 3.80 | 3.60 | 3.40 | 3.20 | 3.00 |
| 17 | 3.80 | 3.80 | 3.60 | 3.20 | 3.00 | 2.80 | 2.60 | 17 | - | - | 4.00 | 3.80 | 3.60 | 3.40 | 3.20 |
| 18 | - | 4.00 | 3.80 | 3.40 | 3.20 | 3.00 | 2.80 | 18 | - | - | 4.00 | 3.80 | 3.60 | 3.60 | 3.40 |

**Figure S3**
PDS youth self-report PDS norms rescaled to 1-4 metric. GAMLSS-estimated percentile curves ($5^{th}$-$95^{th}$) for self-report PDS average scores (range: 1-4) across ages 12-18 years, shown for (A) boys and (B) girls, with (C) the reference percentile table.
